# Multi-Source Graph Synthesis (MUGS) for Pediatric Knowledge Graphs from Electronic Health Records

**DOI:** 10.1101/2024.01.14.24301302

**Authors:** Mengyan Li, Xiaoou Li, Kevin Pan, Alon Geva, Doris Yang, Sara Morini Sweet, Clara-Lea Bonzel, Vidul Ayakulangara Panickan, Xin Xiong, Kenneth Mandl, Tianxi Cai

## Abstract

The wealth of valuable real-world medical data found within Electronic Health Record (EHR) systems is particularly significant in the field of pediatrics, where conventional clinical studies face notably high barriers. However, constructing accurate knowledge graphs from pediatric EHR data is challenging due to its limited content density compared to EHR data for the general population. Additionally, knowledge graphs built from EHR data primarily covering adult patients may not suit the unique biomedical characteristics of pediatric patients. In this research, we introduce a graph transfer learning approach aimed at constructing precise pediatric knowledge graphs. We present MUlti-source Graph Synthesis (MUGS), an algorithm designed to create embeddings for pediatric EHR codes by leveraging information from three distinct sources: (1) pediatric EHR data, (2) EHR data from the general population, and (3) existing hierarchical medical ontology knowledge shared across different patient populations. We break down these code embeddings into shared and unshared components, facilitating the adaptive and robust capture of varying levels of heterogeneity across different medical sites through meticulous hyperparameter tuning. We assessed the quality of these code embeddings in recognizing established relationships among pediatric codes, as curated from credible online sources, pediatric physicians, or GPT. Furthermore, we developed a web API for visualizing pediatric knowledge graphs generated using MUGS embeddings and devised a phenotyping algorithm to identify patients with characteristics similar to a given profile, with a specific focus on pediatric pulmonary hypertension (PH). The MUGS-generated embeddings demonstrated resilience against negative transfer and exhibited superior performance across all three tasks when compared to pediatric-only approaches, multi-site pooling, and semantic-based methods. MUGS embeddings open up new avenues for evidence-based pediatric research utilizing EHR data.

## Introduction

Clinical trials are a well-established method for generating real-world evidence for the study of disease diagnosis and management [1]. Among observational study methods, prospective studies following cohorts and participants’ disease status over extended time periods to identify disease risk factors are the gold standard for establishing causality [2–4]. Not only are clinical trials and prospective studies time and resource intensive, however, but pediatric trials and studies are also particularly difficult due to factors including additional legal regulations, ethical dilemmas, and commercial profitability concerns [1,5,6]. The proportion of clinical drug trials that are pediatric lags significantly behind the pediatric proportion of studied disease burdens [1,5,7–9]. Moreover, many pediatric clinical trials suffer from poor enrollment and high termination rates [6,9,10].

The difficulty and dearth of pediatric trials and studies contribute to a lack of real-world evidence for the pediatric population. Given the persistent shortfall of pediatric research in spite of governmental incentivisation and regulation [11], data stored in electronic health record (EHR) systems provide a valuable alternative source for generating real-world evidence. EHR data in the form of diagnostic and procedure billing codes, laboratory test records, and medication prescriptions push the frontier of clinical research. For example, EHR data have been used to evaluate treatments and patient care models as well as to develop predictive models for diagnosis, treatment, and clinical outcomes [12–22].

The wealth of data in EHR data expands the breadth of biomedical modeling and research, but the scale of the data also poses challenges. A critical step in studying a disease of interest using EHR data is curating a cohort from the clinical population that satisfies specific study criteria. Computable phenotypes are a standardized, machine-readable approach for defining and identifying cases of a disease of interest using its demographic profile, symptoms, laboratory tests, and other clinical information. Given their broad consideration of clinical characteristics, computable phenotypes are also particularly useful in the study and development of precision medicine. Manually selecting or creating the relevant medical features for computable phenotypes from thousands of EHR features is time intensive and requires extensive domain knowledge. For poorly understood diseases, feature selection is even more difficult [23,24]. This has opened another area of EHR clinical research into medical feature extraction and concept representations [25,26], such as knowledge graphs. Algorithmic methods for developing knowledge graphs can efficiently synthesize patient data and existing knowledge.

Several automated and semi-automated methods for generating knowledge graphs have been developed to guide feature selection and modeling strategies [22,27–33]. Many of these knowledge graphs, however, are trained on general population or adult EHR data. Adult knowledge graphs are not directly relevant to or accurate for pediatric populations. Child-specific diseases, treatments, and preventative care create fundamental vocabulary differences between adult and pediatric EHR data [6,34]. Even for shared EHR features, knowledge from the general population may not precisely reflect pediatric disease progression or management. Pediatric patients often require higher standards of precision or different rounding strategies in numeric calculations that are not reflected in adult EHR data or studies [6,34]. Metabolic pathways and rates, receptor functions, and homeostatic mechanisms change from childhood to adulthood. In addition to physiological differences, pharmacological factors such as dosing protocols, drug efficacy and side effects, and therapeutic windows for pediatric patients also differ from that of adult patients [35–38]. The relative importance of features such as social history and caregiver information may also differ between pediatric and general population patients [6,34].

Knowledge graphs specific to children are difficult to generate from pediatric EHR alone, however, due to the relative health of children and resulting sparsity of pediatric EHR. Long before the introduction of EHR systems, pediatric healthcare recordkeeping developed to document well-check visits while the problem-oriented evolution of general healthcare records reflected the general medical care model [39]. Approximately half of pediatric healthcare visits are well-check visits and many developmental screening tests and vaccinations are unique to pediatric patients [6,34]. The preventative care model for children renders pediatric EHR uniquely sparse as compared to EHR data for the general population. As such, existing methods for building knowledge graphs that were developed using general EHR data are ill-equipped to handle the unique sparsity of pediatric EHR data.

To overcome the challenges in constructing pediatric knowledge graphs, we propose a method for transferring knowledge from a general population healthcare system. We draw knowledge from structured EHR data in the form of EHR codes for disease diagnoses, medications, laboratory measurements, and procedures. Relationships between EHR codes, and thus knowledge graphs, can be learned using co-occurrence data of EHR codes and inferred from trained lower dimensional representations, known as embedding vectors, for each code. In spite of the previously outlined differences between pediatric and adult medical data, there are still many shared codes and characteristics between health care systems serving pediatric patients and health care systems serving the general population. This shared foundation forms the basis for synthesizing EHR data and facilitating the transfer of knowledge.

In this study, we introduce the MUlti-source Graph Synthesis (MUGS) algorithm, designed to learn accurate code embeddings from sparse pediatric EHR data. This is achieved through leveraging two additional sources: prior hierarchical medical ontology knowledge and EHR data from the general population obtained from a second information-rich system. By synthesizing these sources, we deconstruct a code embedding into three effect components: the group effect, the overlapping code effect, and the code-site effect. The first two effect types are shared across sites, portraying the homogeneity and similarity. More specifically, the group effect defined by the shared hierarchical medical ontology, has the potential to enhance the transferability of knowledge from the information-rich site. The third effect type is strategically employed to encompass the potential heterogeneity that might exist across sites. Operating within this decomposition framework, our algorithm systematically learns embeddings for each site-specific code via alternating penalized linear regressions. Fine-tuning the hyperparameters in our model not only ensures adaptability to varying degrees of heterogeneity between sites but also fortifies our approach against any adverse effects of negative transfer.

We substantiated the superior performance of MUGS-generated embeddings in comparison to pediatric-only, multi-site pooling and semantic-based methods by their ability to identify established pediatric codes relationships and recognize codes linked to six target diseases. These gold standard code-code pairs for evaluation were curated from reputable online sources, endorsed through pediatric physician input garnered via a survey, or validated using GPT. Moreover, MUGS embeddings enhance the understanding of the heterogeneity between pediatric patients and general patients. In this study, we focused on epilepsy and pulmonary hypertension (PH). They also enable high-quality downstream tasks including the creation of pediatric knowledge graphs, pediatric patient classification and clustering, and more. To provide an accessible avenue for visualizing the pediatric knowledge graphs rooted in MUGS embeddings, we’ve developed an intuitive Shiny App, which can assist pediatric computable phenotyping. Additionally, we formulated a phenotyping algorithm to identify ’patient-like-me’ profiles, concentrating on pediatric PH.

## Results

### Data Preprocessing

We utilized EHR data from Mass General Brigham (MGB) encompassing 2.5 million patients and Boston Children’s Hospital (BCH) encompassing 0.25 million patients in four codified domains: PheCode for diagnoses, RxNorm for medications, Logical Observation Identifiers Names and Code (LOINC) [40] for laboratory measurements, and Clinical Classifications Software (CCS) for procedures. After frequency control, we found that 3055 codes are shared between the two hospital systems, while 1221 codes are unique to BCH and 2350 codes are unique to MGB. The top-down hierarchical structure of medical ontology concepts shared across different hospital systems allows a comprehensive aggregation of the codified data into a common ontology. After applying the hierarchical roll-up, we obtained 337 groups for PheCodes, 304 groups for RxNorm codes, and 700 groups for LOINCs. We then generated a summary-level co-occurrence matrix of EHR codes for each site as described in [25]. The co-occurrence matrix of MGB data is the same as that used in [27]. To allow for further analysis of the dependency between codes, we constructed a shifted positive pointwise mutual information (SPPMI) matrix [41] from each co-occurrence matrix.

### Model Overview: Knowledge Transfer via MUGS

MUGS co-trains embeddings for medical codes using SPPMI matrices from MGB and BCH. Due to the imbalance of EHR data in MGB and BCH, it transfers useful knowledge from the information-rich site MGB to enhance the quality of embeddings in the information-sparse site BCH. Each embedding is decomposed into three components: the group effect, the code effect, and the code-site effect. The group effect incorporates prior knowledge from the hierarchical medical ontology, which is shared across sites and within groups. The code effect represents site nonspecific knowledge shared for a code across sites. The code-site effect depicts site-specific knowledge, crucial for capturing the potential heterogeneity across sites.

To regularize the model, we employ two penalties for the code effect and code-site effect, respectively. When the penalty for code-site effect approaches infinity, it reflects thorough homogeneity between the sites, that is, any overlapping code has the same embedding across sites. When both penalties approach infinity, codes in the same ontology-defined group share one embedding across sites. Conversely, when both penalties are zeros, MUGS degenerates to direct singular value decomposition (SVD) of the SPPMI matrix within each site. Consequently, MUGS is robust against negative transfer and is guaranteed to be no worse than the single-site Skip-Gram method, which is equivalent to conducting SVD of a SPPMI matrix [41]. MUGS can adaptively identify homogeneous codes and heterogeneous codes via a tuning procedure. Details are available in the Supplement. Specifically, as depicted in **Figure 1, Panel (B)**, MUGS assigns the same embedding to codes with similar meanings for general and pediatric patients, such as bacterial enteritis, while assigning different embeddings for codes that perform differently for these patient groups, such as epilepsy.

**Figure 1.**
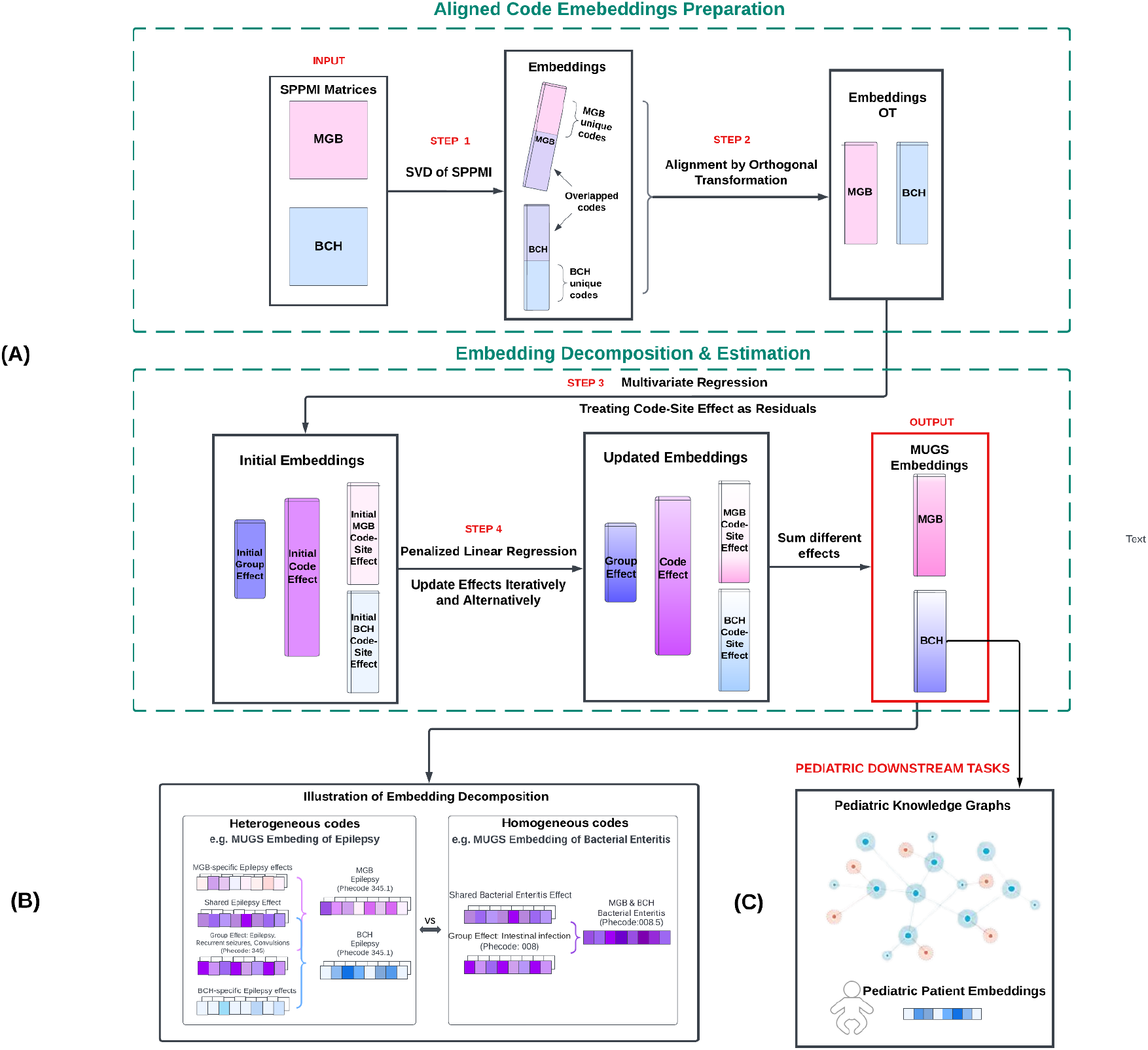
**(A)** Schematic of the overall MUGS algorithm, **(B)** illustration of a heterogeneous code, PheCode:345.1 (Epilepsy), and a homogeneous code, PheCode:008.5 (Bacterial Enteritis), between MGB and BCH, and **(C)** potential pediatric downstreams using MUGS embeddings as input.

MUGS has mian four steps: (1) conducting SVD of SPPMI matrix within each site, (2) performing orthogonal transformation to align multiple sets of embeddings, (3) training initial estimators for group effects, code effects, and code-site effects by pooling aligned embeddings across sites, (4) updating the group effects, code effects, and code-site effects through alternating and iterative solutions of penalized linear regressions until convergence. **Figure 1, Panel (A)** outlines the procedures with MGB and BCH data, and a more detailed and comprehensive explanation is available in Knowledge Transfer via MUGS of Methods Section. As depicted in **Figure 1, Panel (C)**, using MUGS embeddings as input, we can perform a wide range of downstream tasks, such as creating pediatric knowledge graphs, learning pediatric patient embeddings for patient classification and monitoring, and more.

### Accuracy, Adaptivity, and Robustness of MUGS Compared with Benchmark Methods

We compared our MUGS method with four benchmark methods. CODER [42] and SapBert [43] are two pre-trained large language models that solely utilize the descriptions of each medical code from BCH without considering EHR data. Skip-Gram is a single-site word embedding method, which conducts SVD on the SPPMI matrix from BCH [41]. PreTraining aggregates co-occurrence information from MGB and BCH as one cooccurrence matrix by adding up the counts of medical codes and then applies SVD to get embedding of each code. With this method, codes that exist in both systems are assigned the same embedding. The dimension of embedding for Code and SapBert was 768 as used in [42][43][42]. The dimension of embeddings for other methods was set for 500. A detailed discussion on how to select the dimension of embedding is available in the Discussion Section.

As illustrated in **Figure 2**, we validated and evaluated the pediatric embeddings trained using different methods through three primary references: pediatric labels curated from literature, manual curation by pediatric physicians, and curation by GPT-3.5 and GPT-4. Detailed curation procedures are available in Gold Label Curation of Methods Section.

**Figure 2.**
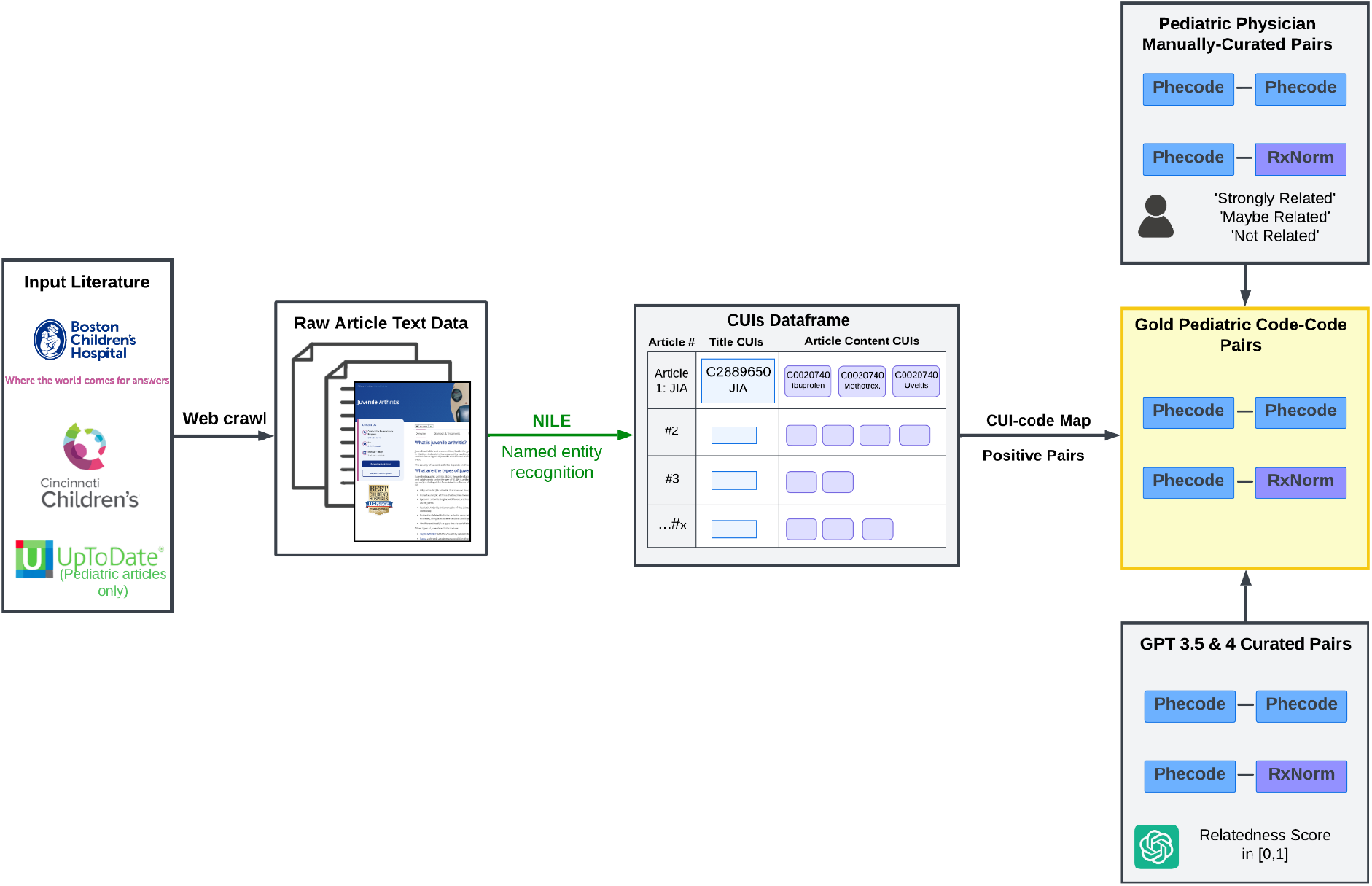
Three primary references on pediatric gold labels for validation and evaluation of pediatric embeddings: semi-automatic positive label curation from literature (BCH website, Cincinnati Children’s Hospital (CCH) website, and UpToDate), manual label curation with three levels by pediatric physicians via a survey on six target diseases, and automatic relatedness score curation by GPT-3.5 and GPT-4 on the six target diseases.

We first assessed the quality of the pediatric embeddings by measuring the accuracy of cosine similarity between two embeddings of each code-code pair in classifying gold standard pairs curated from literature versus random pairs. Gold standard pairs curated from literature indicate the presence of strong relationships, while the sparsity of the network allows us to assume that each random pair represents a weak or no relationship. **Figure 3** summarizes the accuracy of different methods using area under the ROC curve (AUC). We can see that MUGS outperforms all four benchmark methods in terms of AUC for different types of code-code pairs from different expert sources. The poor performance of Coder and SapBert, which were pre-trained based on a medical knowledge graph named the Unified Medical Language System (UMLS) developed for the general patient population, illustrates the heterogeneity between the general patient population and the pediatric patient population. It also suggests that EHR data from BCH is a valuable source for enhancing the quality of pediatric embeddings. Moreover, PreTraining, which leverages EHR data from MGB, beats Skip-Gram, which only utilizes EHR data from BCH. This suggests a degree of similarity between MGB and BCH, enabling knowledge transfer from MGB to improve the quality of pediatric embeddings.

**Figure 3.**
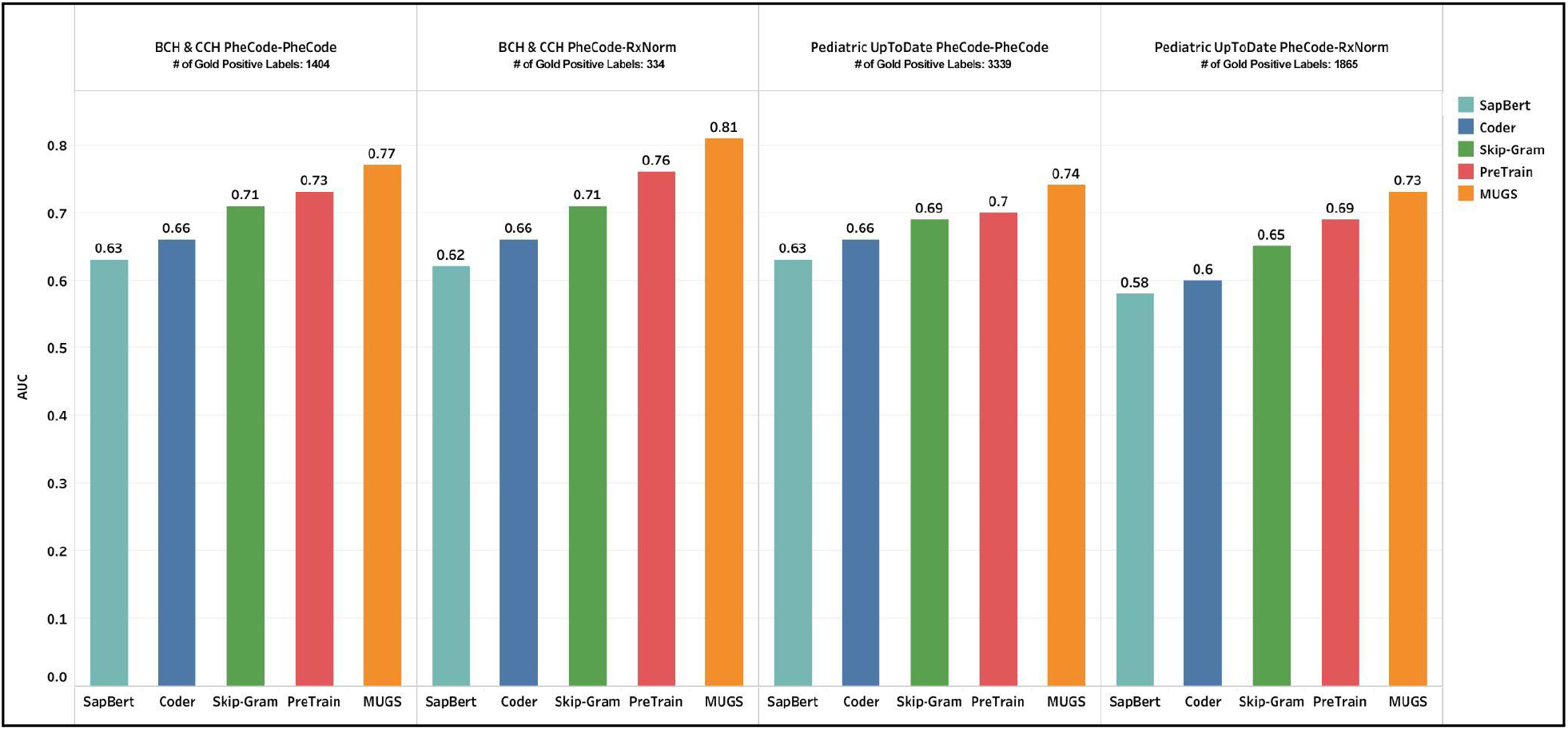
Comparison on AUC between MUGS and benchmark methods using gold labels curated from articles in children’s hospital websites (BCH and CCH) and pediatric articles from UpToDate website.

To further highlight the potential advantages of MUGS, we categorized the codes into two groups based on code frequencies: rare codes, representing those with a patient proportion less than 0.1%; and frequent codes, encompassing those with a patient proportion greater than or equal to 0.1%. By selecting a cutoff of 0.1%, we ensured a sufficient number of rare codes for accurate evaluation within this category, and the marginal frequency of each rare code is relatively low. The accuracy results are summarized in **Figure 4**. Compared with single-site method Skip-Gram, both PreTraing and MUGS exhibit higher accuracies in terms of rare codes by leveraging knowledge from MGB. More importantly, MUGS surpasses PreTraining by enabling the adaptive identification of heterogeneous codes, especially frequent ones, which possess distinct embeddings in MGB and BCH.

**Figure 4.**
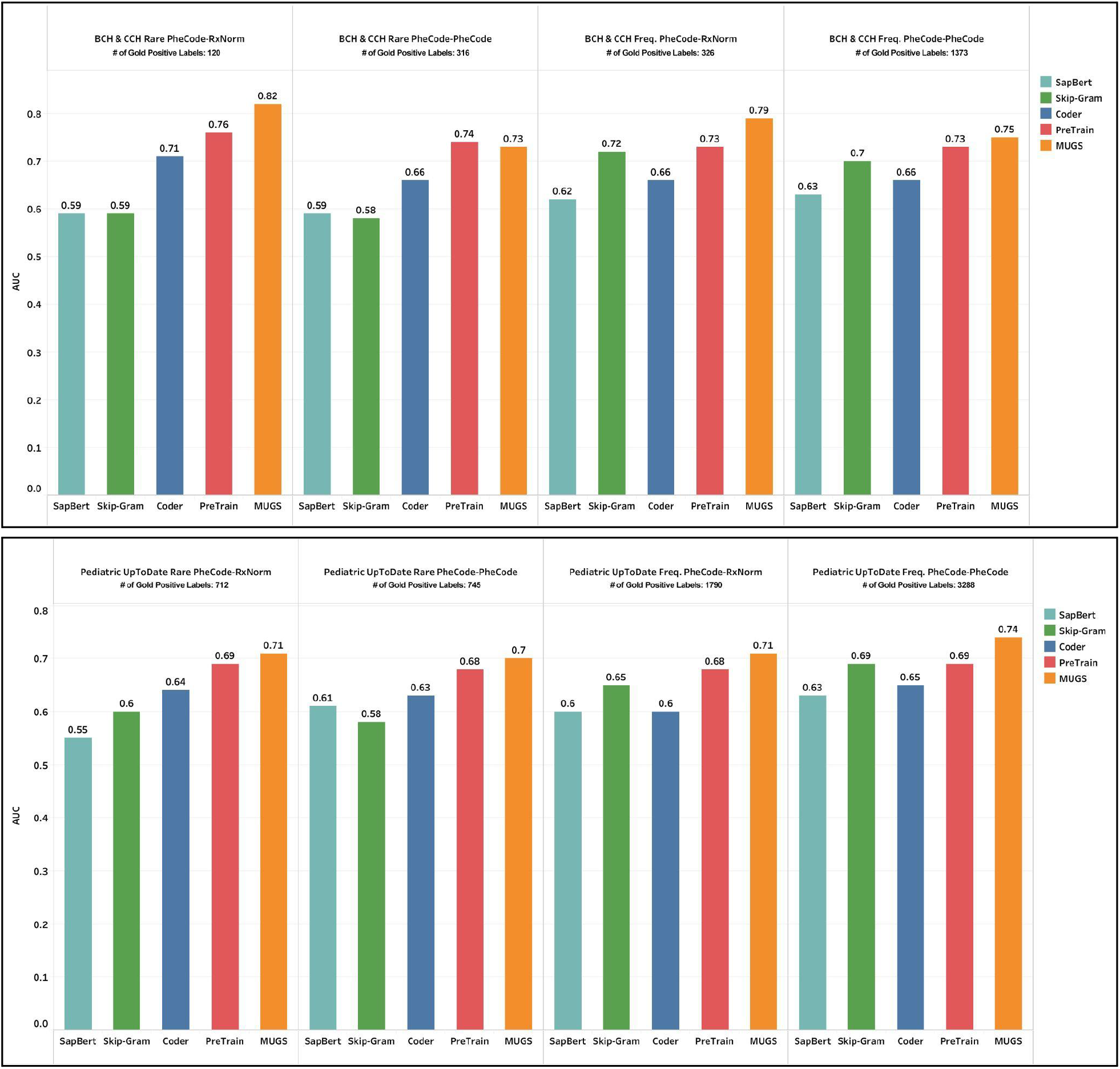
Comparison on AUC between MUGS and benchmark methods using gold labels curated from articles in children’s hospital websites (BCH and CCH) and pediatric articles from UpToDate website. Rare pairs are those in which at least one code is rare, while frequent pairs are those in which at least one code is frequent.

Secondly, we evaluated the quality of the pediatric embeddings by calculating the rank correlation (Kendall’s tau) between cosine similarity, mean scores of gold labels manually curated by pediatric physicians, and ranking scores curated by GPT-3.5 and GPT-4 for six target diseases: Epilepsy, PH, Asthma, Type 1 Diabetes, Ulcerative Colitis, and Crohn’s disease. **Figure 5** clearly demonstrates that MUGS outperforms benchmark methods, exhibiting the highest rank correlations with gold manual labels, GPT-3.5 labels, and GPT-4 labels. Considering that the rank correlation between GPT-3.5 and GPT-4 is 0.81, the relatively high rank correlations of MUGS with the three types of gold labels further verify the quality of MUGS pediatric embeddings. More details on the evaluation methods can be found in Evaluation in Methods Section.

**Figure 5.**
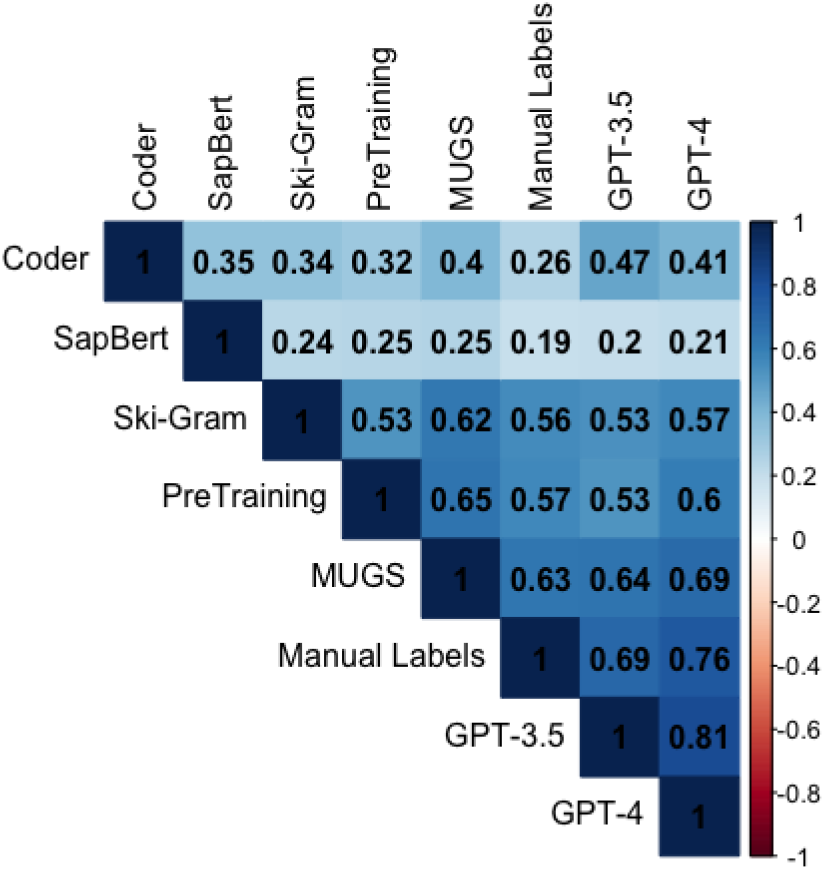
Average Kendall’s tau is computed for six target diseases between different methods. Cosine similarities are used for MUGS and benchmark methods, mean scores between 0 to 1 are used for Manual Labels, and ranking scores between 0 to 1 are used for GPT-3.5 and GPT-4.

### Homogeneity and Heterogeneity in Epilepsy and PH for General and Pediatric Patients

Figure 6. and **Figure 7**. illustrate the relationship among the 15 codes with highest cosine similarity for epilepsy and PH, respectively, and compare the cosine similarity between BCH and MGB. The analysis demonstrates that MUGS effectively captures both the homogeneity and heterogeneity between general patients and pediatric patients with the two diseases. Notably, a total of 13 codes exhibit shared occurrence across both MGB and BCH datasets for epilepsy, and 12 codes demonstrate such shared occurrence for PH in terms of procedures, laboratory tests, conditions including comorbidities and symptoms, and medications, highlighting the substantial similarity present between the two sites regarding the two respective diseases.

MUGS embeddings also effectively capture expected heterogeneity between populations. For instance, infantile cerebral palsy, a group of intellectual and movement disorders that emerge in early childhood, ranks as the ninth related feature concerning epilepsy at BCH, with a cosine similarity of 0.50, while at MGB, it is the 15th-most related feature with respect to epilepsy, with a cosine similarity of 0.41. Topiramate is among the drugs most closely associated with epilepsy at BCH but has lower cosine similarity at MGB, perhaps reflecting the more diverse uses of the drug in adult populations, including prevention of migraines, treatment of psychiatric disorders, and management of weight [44].

**Figure 6.**
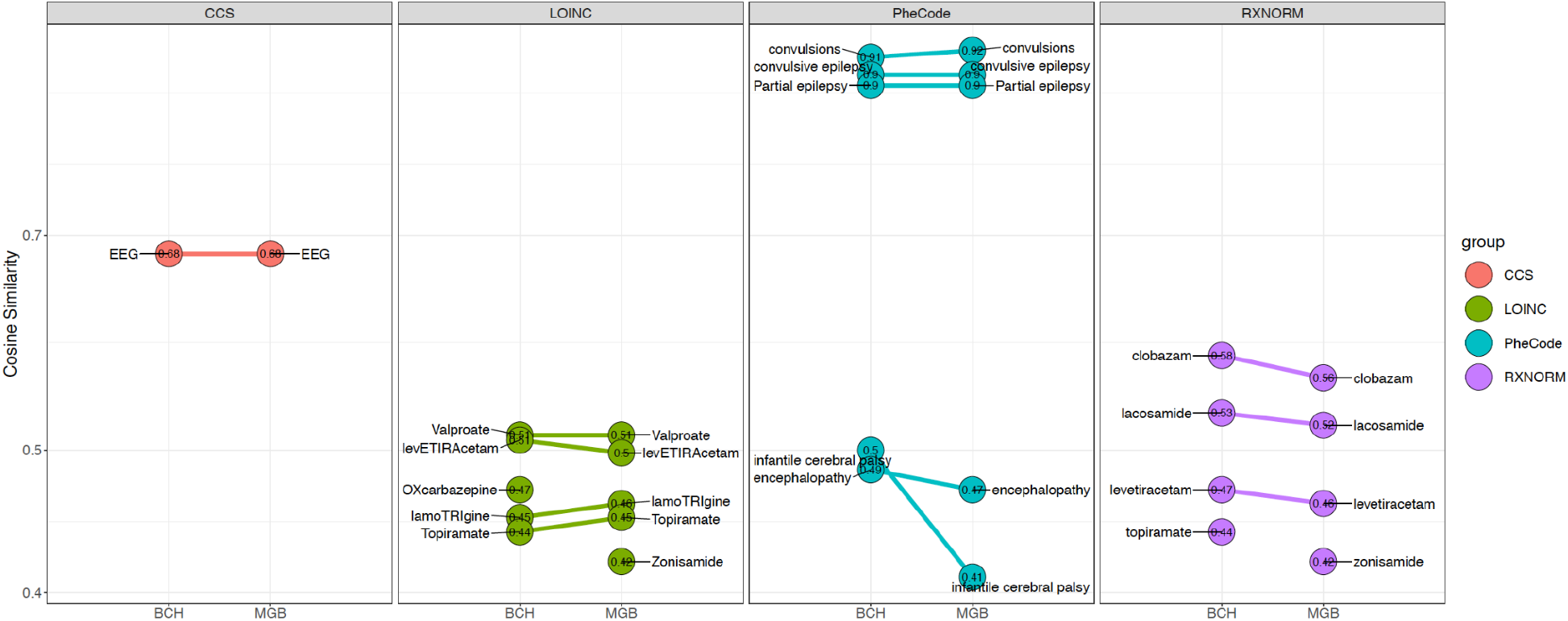
Top 15 codes selected based on the cosine similarity with epilepsy using MUGS embeddings for MGB and BCH. For conciseness, parent PheCodes are omitted if child codes are presented. LOINC codes Valproate, levETIRAcetam, lamoTRIgine, Topiramate, Zonisamide and OXcarbazepine are derived by moving up one layer in the LOINC hierarchy. For example, Topiramate (LP19239-0) is the parent code encompasses various leaf codes, such as topiramate [Mass/volume] in Blood (LOINC: 60192-2), and Topiramate [Mass/volume] in Urine (LOINC: 60193-0).

**Figure 7.**
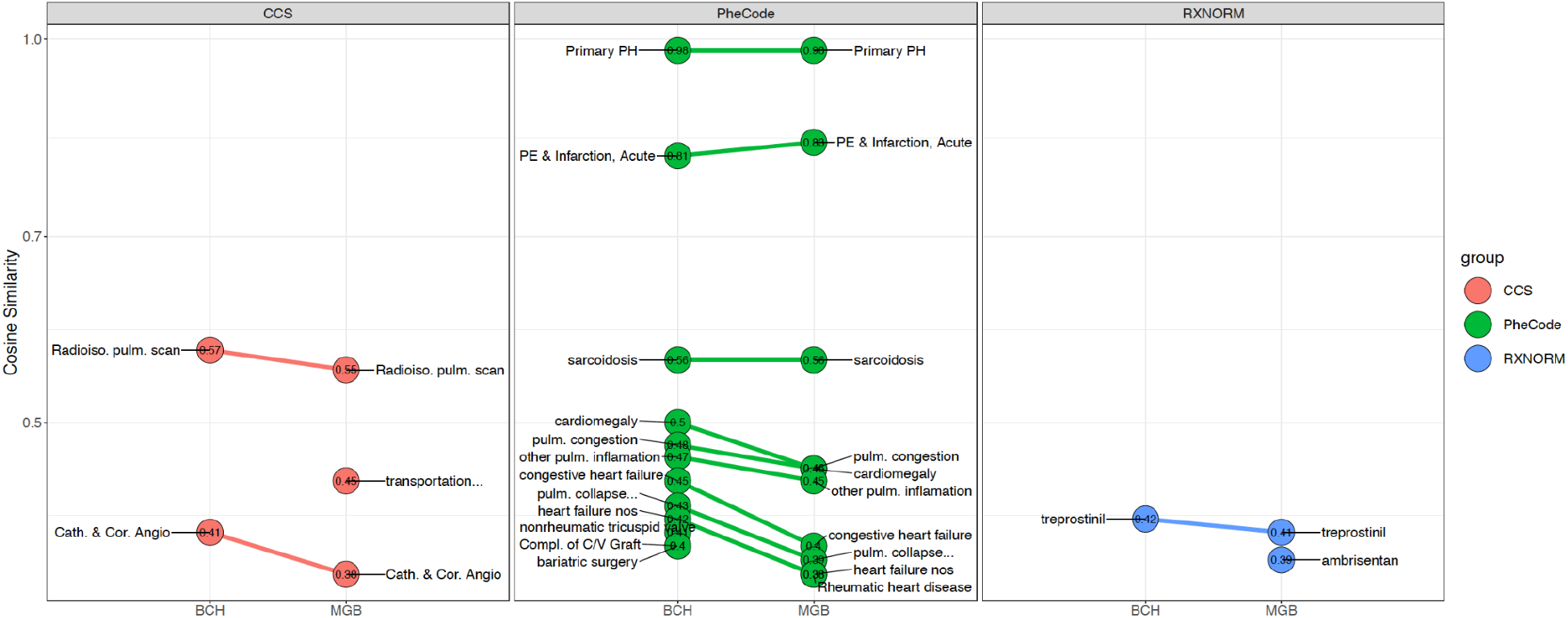
Top 15 codes selected based on the cosine similarity with PH using MUGS embeddings for MGB and BCH.

Shifting to the realm of PH-related features, notable differences between BCH and MGB include rheumatic heart disease being selected only at MGB as an associated condition, while complications of cardiac/vascular device and nonrheumatic tricuspid valve disorders are only selected at BCH. These contrasts reflect expected differences in population where BCH cares primarily for patients with congenital heart disease while MGB cares primarily for patients with acquired heart disease. Similarly, ambrisentan was not selected as a highly associated medication at BCH but was at MGB, reflecting known practice variations between centers.

Another illustration on homogeneity and heterogeneity via knowledge graphs created in a web API with the two diseases between general and pediatric patients can be found in **Figure S1**. and **Figure S2**. in Supplement.

### Epilepsy or PH Related Codes Identification for Pediatric Patients

**Figure 8. and Figure 9**. depict feature clouds generated using embeddings for epilepsy and PH, respectively. The clouds were created using MUGS and Skip-Gram embeddings based on cutoffs of cosine similarity for four categories: PheCode-PheCode, PheCode-RxNorm, PheCode-LOINC, and PheCode-CCS. Detailed information on how these cutoffs are selected is provided in Feature Selection: Cutoff of Cosine Similarity in the Methods section. For epilepsy, with the same cutoff selection method, the number of selected features using Skip-Gram embeddings is 16, compared to 130 using MUGS embeddings, primarily due to the sparsity of BCH EHR data. Consequently, important features such as infantile cerebral palsy and lacosamide, an antiepileptic medication, were not selected by the Skip-Gram method but were captured by our MUGS method. MUGS embeddings also identified detailed CSF testing including CSF amino acid profiling and neurotransmitters, important components of diagnosing epilepsy that are particular to the pediatric population. Similarly, in the context of PH, Skip-Gram embeddings identified 33 related features, whereas MUGS embeddings identified 58. For example, Bosentan, which is primarily used to treat PH, was solely selected by our MUGS method. MUGS also uniquely identified the association between PH and pulmonay embolism. In contrast, the Skip-Gram method identified degeneration of the macula and posterior pole of the retina (PheCode: 362.2) as a relevant condition, but the association between this condition and PH is not readily apparent.

**Figure 8.**
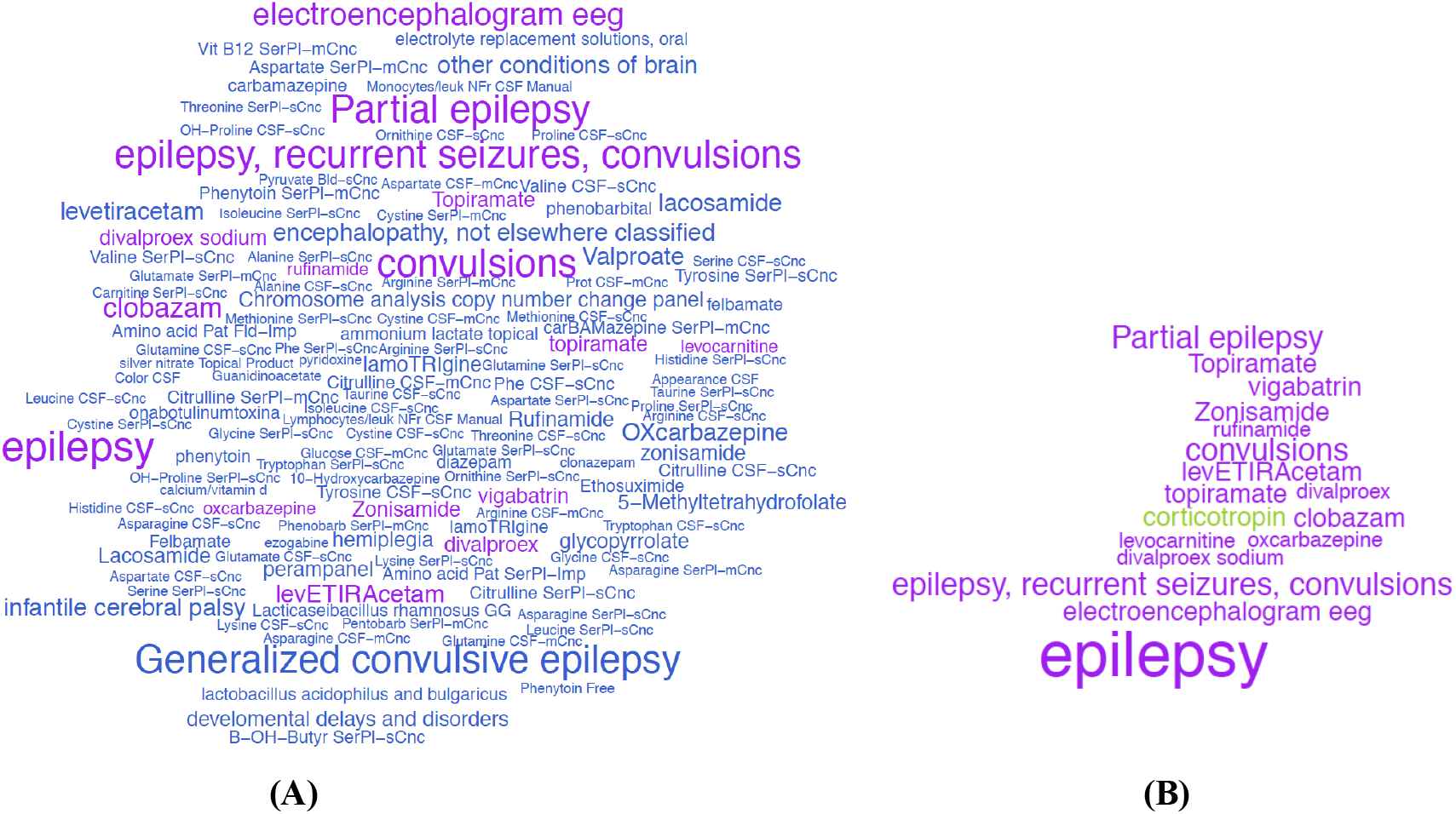
Feature clouds for epilepsy based on cosine similarity of **(A)** MUGS embeddings and **(B)** Skip-Gram embeddings for BCH. Codes selected by both methods are in purple, codes only selected by MUGS are in blue, and codes only selected by Skip-Gram are in green. The size of the words is proportional to the cosine similarity of embeddings of each code-code pair.

**Figure 9.**
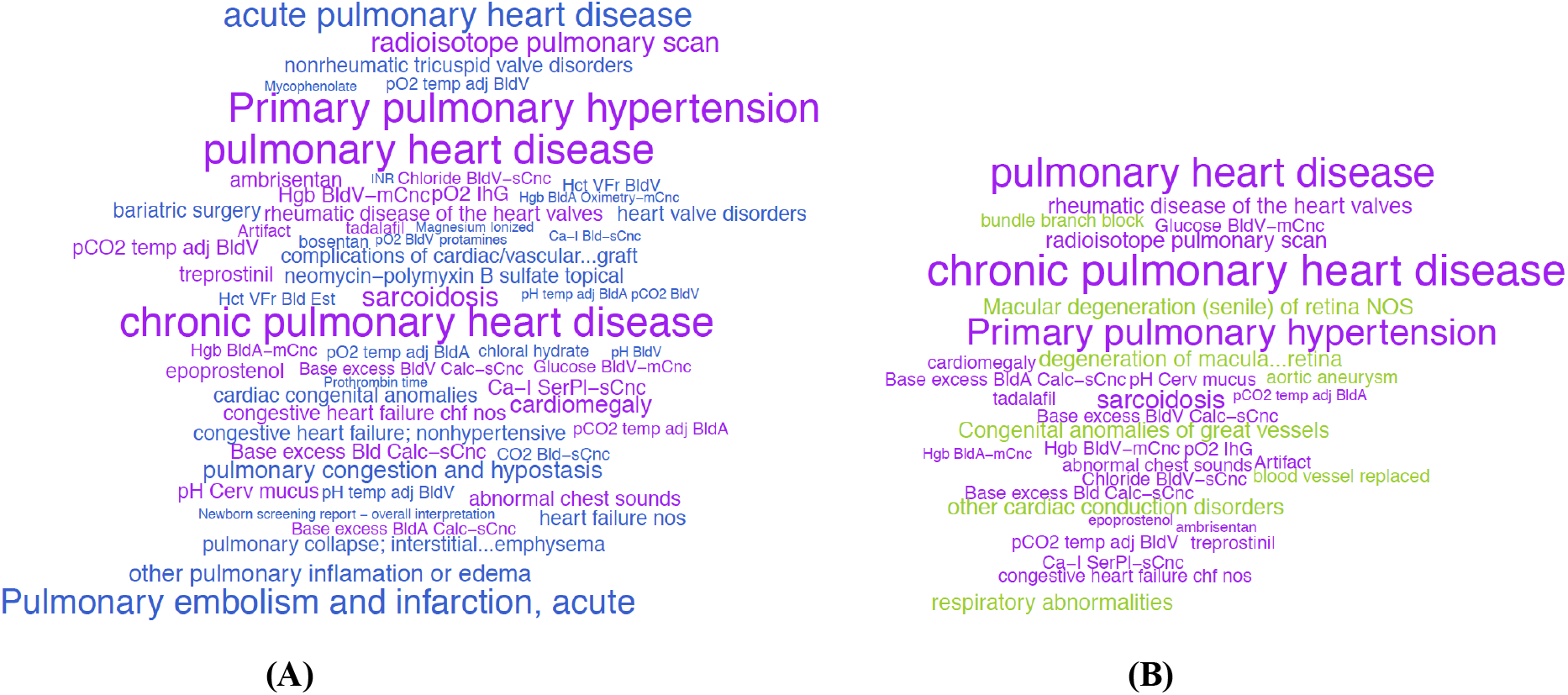
Feature clouds for PH based on cosine similarity of **(A)** MUGS embeddings and **(B)** Skip-Gram embeddings for BCH. Codes selected by both methods are in purple, codes only selected by MUGS are in blue, and codes only selected by Skip-Gram are in green. The size of the words is proportional to the cosine similarity of embeddings of each code-code pair.

### Classifying Pediatric Patients for the Diagnosis of PH

With the target disease PH, we used a dataset comprising 91 pediatric patients from BCH whose records included at least one PheCode:415.2. Each patient’s true PH status was labeled by a domain expert via manual chart review [45]. Among them, 66 were labeled as positive, indicating a diagnosis of PH, and the remaining 25 patients were labeled as absent of PH diagnosis.

The performance of patient classification, utilizing patient embeddings generated from the aforementioned five distinct sets of code embeddings, is summarized in **Table 1**. Additionally, a benchmark method relying solely on the count of ICD code corresponding to PH was employed for comparison. Except the ICD-Benchmark, the five patient-embedding-based methods harness the interconnectedness between various medical codes and PH by gauging the cosine similarity between the respective code embeddings. The detailed patient screening procedure, patient embedding construction, and classification methodology are elaborated in Patient Embeddings Generation and ‘Patient-like-me’ Classification in Methods Section. Furthermore, a visual representation of patient classification with PH using MUGS embedding can be found in **Figure 10**.

**Table 1.** The mean and standard deviation (SD) of 150 AUCs, sensitivities and positive predictive values (PPV) at specificity = 0.95 on classifying pediatric patients with and without PH of five different code embeddings generating methods, alongside an ICD-Benchmark (ICD) method soly utilizing the count of PheCode:415.2.

|  | ICD | Coder | SapBert | Skip-Gram | PreTraining | MUGS |
| --- | --- | --- | --- | --- | --- | --- |
| AUC | 0.941 (0.038) | 0.698 (0.084) | 0.710 (0.086) | 0.951 (0.032) | 0.924 (0.047) | **0.961 (0.030)** |
| Sensitivity | 0.772 (0.126) | 0.216 (0.181) | 0.248 (0.208) | 0.801 (**0.111**) | 0.679 (0.169) | **0.822 (0.115)** |
| PPV | 0.975 (0.004) | 0.819 (0.225) | 0.871 (0.238) | 0.976 (0.004) | 0.971 (0.009) | **0.977 (0.004)** |

**Figure 10.**
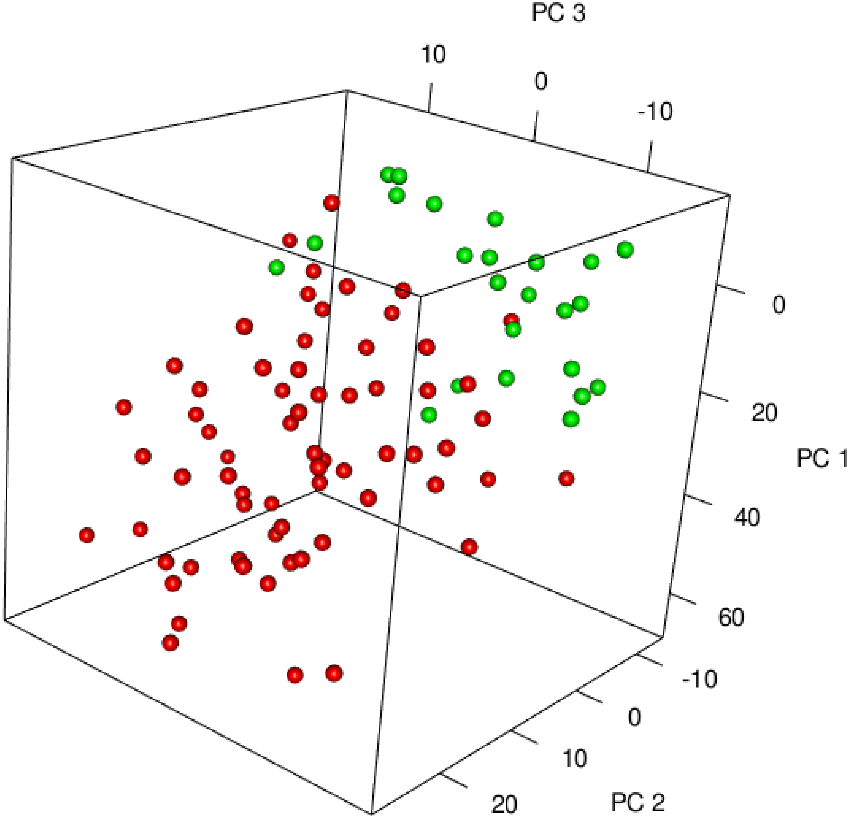
Three-dimensional illustration of pediatric patients classification with PH based on the first three principle components (PCs) of MUGS embeddings for BCH. In the figure, red points represent patients diagnosed with PH, while green points represent patients without a PH diagnosis.

Notably, Coder and SapBert are significantly worse than other methods which utilize EHR data in classifying pediatric patients for PH diagnosis. This stark contrast underscores the substantial value of leveraging EHR data for accurate patient classification and the identification of ’patient-like-me’ profiles. MUGS and Skip-Gram outperform ICD-Benchmark, exhibiting the efficiency gained through capitalizing on the relatedness between medical codes learned with EHR data. Comparatively, the performance of PreTraining is worse than that of the ICD-benchmark and Skip-Gram, which indicates significant heterogeneity in PH presentation between pediatric patients and general patients. MUGS emerges as the most effective method with the highest AUC, sensitivity, and positive predictive values (PPV) at specificity = 0.95, which further illustrates its efficiency, adaptivity and robustness in code embeddings construction and consequently, patient classification.

## Discussion

The MUGS approach efficiently and robustly summarizes patient-level EHR data into institution-specific code embeddings by leveraging existing hierarchical medical ontologies and co-training EHR data from multiple institutions. The novel integration of these additional data sources empowers the generation of high-quality embeddings, even from sparse EHR data, adaptively capturing the inherent heterogeneity across EHR populations. The MUGS approach thus facilitates knowledge graph construction, feature extraction, computable phenotyping, and patient clustering in studying pediatric patients and other populations with sparse EHR data. MUGS outperforms existing methods in several aspects. First, it enhances knowledge transfer across sites via a shared hierarchical medical ontology. Second, it can adaptively capture the homogeneous codes and heterogeneous codes across sites as well as the strength of different group effects through hyperparameter tuning. Last, it maintains robustness against negative transfer, ensuring no performance degradation when introducing data from another site.

MUGS embeddings can be used for several downstream tasks that rely on accurate concept embeddings tailored to unique patient populations. This becomes especially pivotal when dealing with information-scarce populations or sites, and it is critical in studying rare diseases and diseases that manifest or progress differently across patient populations. An essential application involves automated feature selection, a process utilizing code embeddings for identifying relevant concepts for studying a disease condition. This can range from straightforward cosine similarity cutoffs to more complex feature selection algorithms. Effective feature selection plays an important role in the construction of knowledge graphs and phenotyping efforts. Furthermore, MUGS embeddings can be used to construct high quality population-specific patient embeddings, facilitating the precise and unbiased development of computable phenotypes, curation of study cohorts and ‘patient-like-me’ identification tailored to specific populations, such as pediatric patients with PH from BCH. The joint representation learning framework adopted in MUGS entails unified embeddings across different institutions, which also allows us to use a specific patient cohort from a “seed” institution, say a small number of PH patients assembled at BCH, to identify “patient-like-me” from another institution.

While the training of MUGS using BCH and MGB data already achieved strong performance results compared to existing methods, there are a few aspects of this study can be improved. One limitation is that MUGS forces the dimension of embedding to be the same across different sites. However, it is possible that the optimal dimension in different sites may vary. In practice, we can use the largest one as the dimension of embedding of MUGS. One potential future work to address this limitation is to allow different dimensions for group effect, code effect and code-site effect in different sites, then concatenate them as a longer embedding for a code in a site. Incorporating code-site weights based on the frequencies of different codes across sites, available in EHR data, is another future work worth studying. It could potentially improve the efficiency of identifying heterogeneous codes, especially frequent and heterogeneous ones. Additionally, the evaluation and validation currently rely on a limited set of gold pediatric labels focusing on PheCode-PheCode and PheCode-RxNorm relationships, which could be expanded to include more relationship types, such as disease and laboratory test pairs or disease and procedure pairs. It could further enhance the assessment of performance of pediatric embeddings. Moreover, when identifying epilepsy and PH related codes for pediatric patients, we used an ad-hoc quantile-based cutoff selection method for feature extraction. Other systematic feature extraction methods, such as sparse embedding regression [27], can also be employed, which might offer additional insights and potential improvements in the identification process. To generate more comprehensive knowledge graphs, unstructured/uncoded concepts from clinical notes can be included as Concept Unique Identifiers (CUIs) when constructing the co-occurrence matrices.

In conclusion, we have demonstrated how MUGS excels in versatile and robust knowledge graph co-training/transfer learning. It co-trains EHR data from MGB and BCH, and effectively transfers knowledge from general patients to pediatric patients, overcoming the challenges of heterogeneity between the populations and sparse pediatric EHR data. MUGS not only facilitates research on understudied pediatric populations but also provides a transfer learning framework for diverse healthcare populations.

## Methods

### Data Preprocessing

BCH is a quaternary referral center for pediatric care and also serves as a primary and specialty care site for the local community. MGB is a Boston-based non-profit hospital system serving primarily an adult population, although MGB also provides neonatal and some general and subspecialty pediatric care. A total of 250,000 patients from BCH and a total of 2.5 million patients from MGB with codified data with at least one visit were included in this analysis.

We gathered four domains of codified data including diagnosis, medication, lab measurements and procedures from BCH and MGB. Diagnosis International Classification of Disease (ICD) codes representing the same general diagnosis were aggregated under a single representative one-digit level PheCode using the ICD-to-Phecode mapping from the PheWAS catalog (https://phewascatalog.org/phecodes) [46]. Local medication codes were consolidated under the RxNorm codes [47]. Local laboratory measurement codes were aggregated under the corresponding LOINC codes. Due to the difference in coding systems between MGB and BCH, we rolled up LOINC codes with the same meaning as a new code. For example, we rolled up LOINC:30341-2 (ESR Bld Qn) and LOINC:4537-7 (ESR Bld Qn Westrgrn) to LP16409-2 (Erythrocyte sedimentation rate). We assigned CCS categories to procedure codes, including CPT-4 (Current Procedural Terminology), HCPCS, ICD-9-PCS, and ICD-10-PCS codes, using the CCS mapping software (https://www.hcup-us.ahrq.gov/toolssoftware/ccs_svcsproc/ccssvcproc.jsp). The top-down hierarchical structures of PheCodes, RxNorm codes, and LOINC codes, allow a comprehensive aggregation of the codified data into a common ontology.

We generated a summary-level co-occurrence matrix of EHR codes for each site as described in [25]. To start, we created an individual co-occurrence matrix of EHR codes for each patient’s extracted data. The rows and columns of these square matrices represent EHR codes, with entries indicating the number of times each pair of codes co-occur in the patient’s EHR record within 30 days. We sumed the individual co-occurrence matrices for all patients in each site to yield a summary-level co-occurrence matrix. If the co-occurrence number is less than 200, we replaced it with zero and removed columns and rows with all zeros from the co-occurrence matrix to achieve frequency control. To further study the dependency between two codes, we constructed the SPPMI matrix for BCH and MGB.

We denote the co-occurrence matrix in site *k* as *C*^(*k*)^, *k* ∈ {1,…,*K*}. In this analysis, we have two sites BCH and MGB, and *K* = 2. We use 𝒱_*k*_ to denote the vocabulary, the set of all medical codes, of site *k*, and *n*_*k*_ to denote the cardinality of 𝒱_*k*_. For the *k*th site, the (*i, j*) th entry of the SPPMI matrix is obtained as 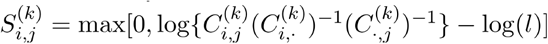, where *l* is the negative sampling value which is often set to 1, i.e., no shifting, 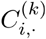 and 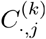 are the *i*th row sum and *j*th column sum of the cooccurrence matrix *C*^(*k*)^, respectively.

### Knowledge Transfer via MUGS

#### Embedding Decomposition

The proposed MUGS algorithm utilizes SPPMI matrices from multiple sites and incorporates shared hierarchical medical ontology knowledge by decomposing the embedding for a code at a given site into group effect, code effect, and code-site effect component embeddings. Formally, we decompose the *p*-dimensional embedding vector for code *i* in site *k*, denoted by u_*ik*_, as

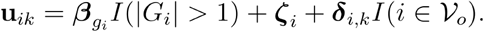

In the first term of the decomposition, index *g*_*i*_ ∈ {1,…, *m*} denotes the group of code *i*, defined by the hierarchical medical ontology, *G*_*i*_ is the corresponding group containing ∣ *G*_*i*_ ∣ codes, and *β*_*gi*_ is the embedding of the effect of group *gi*. In the second term, *ζ*_*i*_ is the effect of code *i*. The third term *δ*_*I*,*k*_ is the code-site effect of code *i* at site *k* that can capture heterogeneity across sites, and 𝒱 _º_ is the set of overlapping codes present in at least two sites, with cardinality *n*. Note that if a group only contains one code, then the group effect and code effect are not separable. Similarly, for non-overlapping codes that only exist in one site, code effect and code-site effect are not separable. To ensure model identifiability, we introduce the indicator function *I*(·) and assume the group effect is non-zero only if the code is part of a nontrivial group (i.e., a group consisting of two or more codes), and code-site effect is non-zero only if the code existing in at least two sites.

#### Penalized Loss Function

The MUGS algorithm learns *β*_*gi*_, *ζ*_*i*_, and *δ*_*i*,*k*_ for all codes across sites from the SPPMI matrices using the following model:

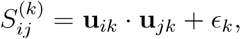

where the error term *∈*_*k*_ represents the noises of SPPMI matrix observed in site *k*. To estimate the group, code, and code-site effects 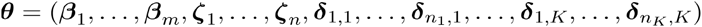, we employ a loss function with penalties for the code and code-site effects:

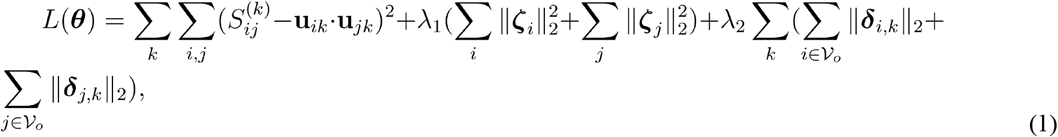

where λ_1_ and λ_2_ are two tunable hyperparameters. We highlight that the hyperparameters λ_1_ and λ_2_ can be tuned adaptively to the degree of heterogeneity across sites and the importance of the hierarchical medical ontology. We can see this by considering the following extreme cases. When the underlying populations of the *K* sites share little similarity, both λ_1_ and λ_2_ would be selected as zeros. Consequently, the penalty terms disappear and the method reduces to direct matrix factorization with respect to SPPMI matrix within each site. In this case, no knowledge will be transferred. When the underlying populations of the *K* sites are exactly the same, λ_2_ would approach infinity. Consequently, the code-site effects would be shrunk to zeros, and the resulting embeddings are dominated by the group and code effects that are shared across sites. When the underlying populations of the *K* sites are exactly the same and codes in a group also act in the same way, both λ_1_ and λ_2_ would approach infinity. In this case, both code and code-site effects are shrunk towards zeros and the resulting embeddings are dominated by group effects. In the analysis with BCH and MGB data, we observed that some codes behave similarly within both pediatric and adult patients, while others exhibit distinct patterns. Certain groups of codes act as cohesive units, while other groups show variability in their individual code behaviors. By tuning λ_1_ and λ_2_ carefully, we can effectively identify homogeneous and heterogeneous codes, and control the signal strength of different group effects.

Additionally, we prioritize the code-site effect in the optimization process, which is the key of capturing potential heterogeneity across sites. With the unsquared L2 code-site penalty, code-site effects are allowed to be shrunk to exact zero, implying these codes are homogeneous across sites, whereas the penalty on code effects is a ridge penalty that prevents overfitting of code effects and controls the signal strength of group effects. The design of our loss function thus ensures adaptability and a robust performance that is no worse than the baseline single-site matrix factorization method, Skip-Gram method.

The decomposed, non-convex nature of the loss function (1) prevents the use of standard optimization algorithms. We propose an alternating and iterative method to minimize it within a broader context, where matrix *S*^(*k*)^ can be asymmetric, which is frequently encountered in recommendation system problems. The detailed procedures on minimization and hyperparameters tuning are given in the Supplement.

### Gold Label Curation

#### Label Curation from Literature

To evaluate the quality of the MUGS embeddings specifically on the pediatric population, we semi-automatically curated new pediatric gold labels by performing named entity recognition on disease-specific articles from three expert sources: BCH website (https://www.childrenshospital.org/conditions), CCH website (https://www.cincinnatichildrens.org/search/health-library), and UpToDate (https://www.wolterskluwer.com/en/solutions/uptodate). The web pages serve as the authoritative source, bypassing the need for individual expert curation. Our process begined by using a web crawler to gather paragraphs of information from each disease page on the websites. For UpToDate, which contains articles about a variety of medical subjects not specific to pediatrics, we only selected articles whose titles contain terms like “child”, “neonate”, or “infant”. Then, we used the Narrative Informative Linear Extraction (NILE) algorithm [48] to identify key CUIs representing diseases and corresponding conditions, symptoms, and treatments. For each disease, we generated CUI-CUI pairs of two general forms: disease-condition and disease-treatment. The CUI pairs were translated into two types of medical code pairs, PheCode - PheCode and PheCode - RxNorm, utilizing an industry-standard CUI-code dictionary, thereby completing the curation of gold pediatric code-code pairs. In total, 13585 relatedness pairs specific to pediatric conditions are created - 5911 pairs of diseases and drugs (PheCode - RxNorm) and 7674 pairs of diseases and conditions (PheCode - PheCode). Of the 13585 pairs created, 798 are from CCH, 1751 are from BCH, and 11036 are from Up-To-Date. Note that not all codes curated here exist in the co-occurrences matrix from BCH.

#### Manual Label Curation by Pediatric Physicians

A survey was conducted among pediatric physicians, focusing on six target diseases: Epilepsy,PH, Asthma, Type 1 Diabetes, Ulcerative Colitis, and Crohn’s disease. For each target disease, a random selection of 10 additional conditions and 10 medications was made. Respondents were asked to indicate their perception of the relationship by choosing from ’strongly related’, ’maybe related’, or ’not related’. We received 14 responses for Epilepsy, 6 responses for Pulmonary Hypertension, and one response each for the remaining four diseases.

#### Label Curation from ChatGPT and GPT4

For each target disease in the survey above, we tasked GPT-3.5 and GPT-4 as AI models with medical knowledge to assign a score on the degree of relatedness between the target disease and each of the 20 medical codes paired with the target disease in the survey. The specific instruction we provided to GPT-3.5 and GPT-4 was as follows: ‘As an AI with medical knowledge, your task is to evaluate the degree of relatedness between two clinical concepts. The objective is to aid in feature selection specific for pediatric patients, implying that the concepts should ideally bear some clinical or medical connection. Please provide your evaluation as a numerical value, rounded to two decimal points, ranging from 0 (no correlation) to 1 (highly correlated). Note: Only respond with a SINGLE numerical value, NO textual explanations.’

### Evaluation

We first assessed the quality of the embeddings by measuring the accuracy in detecting gold standard pairs (PheCode-PheCode and PheCode-RxNorm) curated from pediatric literature. Within each type of relation, majority of the pairs are unrelated. The sparsity of the network allows us to assume that each random pair represents a weak or no relationship. For each type of relation, using the gold labels curated from a certain source of the literature, we randomly generated an equal number of random pairs, created a binary vector of 1’s and 0’s indicating the presence or absence of a strong relationship. We then computed the cosine similarity between the embeddings in each pair. The accuracy was summarized using the AUC between the cosine similarity and the binary vector.

For manual labels curated by physicians, we recoded ’strongly related’ as 1, ’maybe related’ as 0.5, and ’not related’ as 0, then calculated the mean score for each pair. For each target disease, labels curated from GPT-3.5 and GPT-4 provided ranking scores between zero and one, reflecting the degree of relatedness. Unlike binary gold labels curated from literature, these ordinal labels contain more information. As a result, we utilized Kendall’s tau as the evaluation criterion instead of AUC. Specifically, for each target disease, we calculated cosine similarity of each pairs using embeddings from four benchmark methods and MUGS. We then computed Kendall’s tau among cosine similarities, mean score from manual labels and ranking score from GPT labels. The average Kendall’s tau of the six target diseases served as the final evaluation criterion.

### Feature Selection: Cutoff of Cosine Similarity

When using a PheCode as the target, we consider four types of pairs: PheCode-PheCode, PheCode-RxNorm, PheCode-LOINC, and PheCode-CCS. To determine suitable cutoffs for feature selection, we randomly generated 5000 pairs for each type and calculated the 99th quantile of their cosine similarities based on code embeddings. We then used these four quantiles as cutoffs to select codes. Specifically, we retained codes whose cosine similarities with the target code were greater than or equal to the respective cutoff within each group.

### Patient Embeddings Generation and ‘Patient-like-me’ Classification

With target disease PH (PheCode:415.2), we selected BCH patients who have received at least one PheCode:415.2 in the corresponding EHR to form the study cohort. After patient screening, we obtained a dataset comprising 2735 patients. Among these patients, 2644 patients did not have any labels related to PH, while 91 patients had labels assigned by pediatric physicians.

We formulated a patient embedding as the weighted sum of code embeddings. The weight assigned to each code for a specific patient is determined by taking the logarithm of one plus the count of times the code appears in the patient’s EHR, divided by the logarithm of one plus the count of patients who received this code. This weight is then further multiplied by the cosine similarity between the code and the target disease calculated using code emebddings.

We assume that the patient embeddings of the case group and the control group follow Gaussian distributions with different means but the same covariance matrix. By minimizing the within-class variance and maximizing the between-class variance, we can identify ‘patient-like-me’. Specifically, to reduce the dimension, we first performed principal component analysis (PCA) on patient embeddings derived from various sets of code embeddings, including Coder, SapBert, Skip-Gram, PreTraining, and MUGS, totaling 2735 labeled and unlabeled patients. We selected the first *M* PCs as predictors and applied them to conduct regularized discriminant analysis (RDA) [49] using labeled data. There are two regularization parameters in RDA. We set the first regularization parameter to one, which yields linear discriminant analysis (LDA). LDA maximizes the ratio of between-class variance to within-class variance. Due to the relatively small labeled dataset, LDA can be poorly-posed. To stabilize the computation, we treated the second regularization parameter in RDA, which controls the shrinkage toward the identity matrix, as a tuning parameter. We also treated *M* as a tunning parameter. We fine-tuned the two tuning parameters through two-fold cross-validation using mean squared error (MSE) as the guiding criterion. We then refitted the model using the selected optimal tuning parameters corresponding to the smallest MSE.

To assess the performance of diverse patient embeddings, we partitioned the data randomly into three folds, employing two for model training and the remaining one for evaluation. To mitigate the impact of random sampling, we repeated this data splitting process 50 times and reported the mean and standard deviation of 150 AUCs, sensitivities, and PPVs as our final outcomes. In addition to the aforementioned five methods, we established another ICD-benchmark outcome. This benchmark employs the logarithm of one plus the count of PheCode:415.2 for each screened patient as a predictor and fits a LDA using labeled data.

## Data Availability

All data produced in the present study are available upon reasonable request to Boston Children's Hospital and Mass General Brigham.

## Supplement

### Optimization Algorithm

Although in this paper, we focus on SPPMI matrices from EHR data, our MUGS approach can be applied to broader settings where *S*^(*k*)^ is asymmetric. For example, 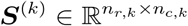 can be a utility matrix of *k*th recommendation system, with *n*_*r*,*k*_ users and *n*_*c*,*k*_ items. Hence, we consider the following more general model:

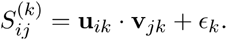

Embedding vector v_*jk*_ can be similarly decomposed into 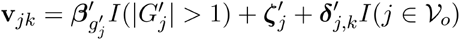, where 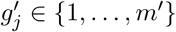 denotes the group of item *j*, and 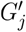 is the corresponding group containing 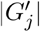 items. Let 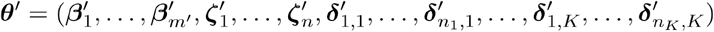, the loss function (1) can be correspondingly extended to

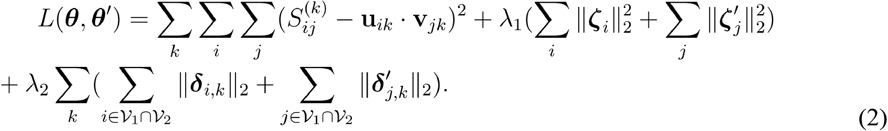

In order to optimize to the correct minimum of the non-convex loss function (2), we first need to find relatively good initial estimators for the effects of interest. The following **Steps 1-3** are on constructing initial estimators of *θ* and *θ′*, while **Step 4** is on updating these effects alternatingly and iteratively until convergence.

**Step 1:** Perform SVD on the SPPMI/utility matrix in each site, and select the top *p* singular values and corresponding *p* left and right singular vectors. Let 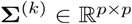 be a diagonal matrix whose diagonal elements consist of these top *p* singular values, arranged in descending order. Let 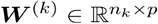 represent the matrix containing the corresponding *p* left singular vectors, and let 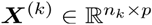 represent the matrix containing the corresponding *p* right singular vectors in in site *k*. The initial Skip-Gram embedding for each code/user and item in site *k* is denoted by 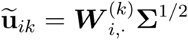 and 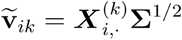, where 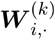 and 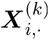 are the *i*th rows of matrices *W*^(*k*)^ and *X*^(*k*)^, respectively.

**Step 2:** Align the directions of *K* sets of initial Skip-Gram embeddings via orthogonal transformation using overlapping codes. Let 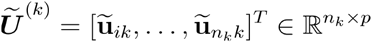 denote the initial Skip-Gram embedding matrix of site *k*, and 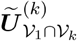 denote the initial Skip-Gram embedding matrix in site *k* of the overlapping codes in site 1 and site *k*. The estimated orthogonal transformation matrix can be obtained as 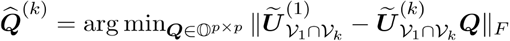, for *k* = 2,…, *K*, where 𝕆^*p*×*p*^ is the set of all *p* × *p* orthogonal matrices, and | | · | |_*F*_ denotes the Frobenius norm. This can be solved by an orthogonal procrustes problem. Then the aligned embedding matrices in site *k* are defined as 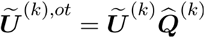 and 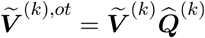, for 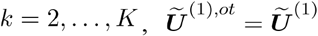, and 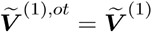. Since 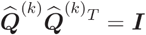, then 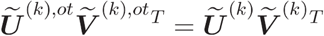.

**Step 3:** Calculate initial estimators for group, code, and code-site effects, denoted by 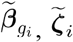, and 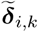, via pooling the aligned initial Skip-Gram embeddings across sites. Specifically, 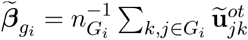, where 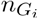 is the sum of number of codes in group *G*_*i*_ in *K* sites, 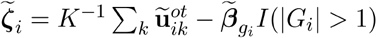, and 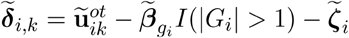, for 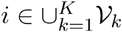 and *k* = 1,…, *K*. For symmetric 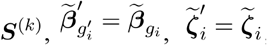, and 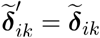. Otherwise, 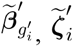, and 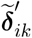 can be estimated similarly using 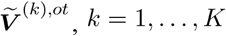.

**Step 4:** For each iteration *t*, update *θ* and *θ′* in an alternating fashion.

**Step 4.1:** Iteratively update 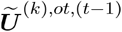, component-wise, by fixing 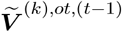:

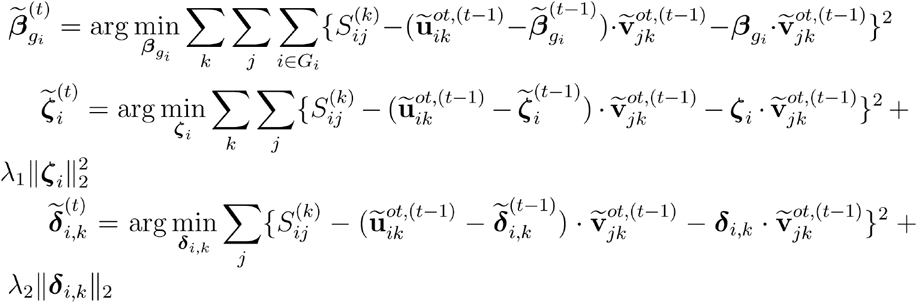

When the stopping condition 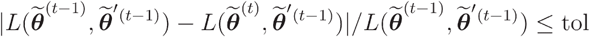, where is the pre specified tolerance, is met, set 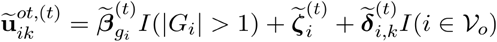.

**Step 4.2:** Iteratively update 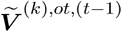, component-wise, by fixing the updated 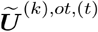. In the same manner as Step 4.1, sequentially and iteratively update 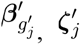, and 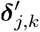 until the stopping condition is met. Then, set 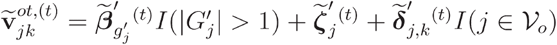.

**Step 4.3:** Repeat Step 4.1 and Step 4.2 until the stopping condition

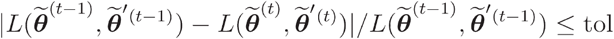

is met. Output 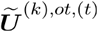 and 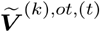 as the final MUGS embeddings.

### Tuning Procedure

We randomly selected 25% gold standard pairs from UpToDate pairs to tune (λ_1,_ λ_2_), and the remaining 75% of gold standard pairs were utilized to assess the quality of the embeddings. During the tuning procedure, we randomly selected an equal number of random pairs as controls. For embeddings trained with a given (λ_1,_ λ_2_), AUC of the cosine similarity in distinguishing gold standard pairs from random pairs was calculated using the 25% gold standard pairs from UpToDate. We select (λ_1,_ λ_2_) maximizing the AUC as the final hyperparameters.

### Multi-View Knowledge Graphs

We have developed a Shiny App (https://dev.parse-health.org/shiny/multi-view-net/) that showcases the knowledge graphs generated using MUGS embeddings for both MGB and BCH. The App offers users the flexibility to utilize MGB-only, BCH-only, or both MGB and BCH MUGS embeddings. It allows users to select one or more target medical concepts and perfrom feature selection by specifying cosine similarity cutoffs for four types of codes in MGB and BCH or determining the number of top nodes to display based on five different sorting criteria.

**Figure S1**, a screenshot of the App, presents a knowledge graph on epilepsy. We identified the top 30 codes most relevant to epilepsy based on the mean of cosine similarities for MGB and BCH with MUGS embeddings. Among these codes, 23 are shared between the two patient populations, denoted by yellow dashed edges, while seven codes are specific to pediatric patients from BCH, indicated by blue dashed edges. For example, one of the specific codes for pediatric patients is Vigabatrin, a medication utilized in managing infantile spasms and refractory complex partial seizures. We also identified the top 30 codes most relevant to PH based on the mean of cosine similarities for MGB and BCH with MUGS embeddings, presented in **Figure S2**. Among these codes, 23 are shared between the two patient populations, while four codes are specific to BCH patients, and three codes are specific to MGB patients, indicated by red dashed edges.

**Figure S1.**
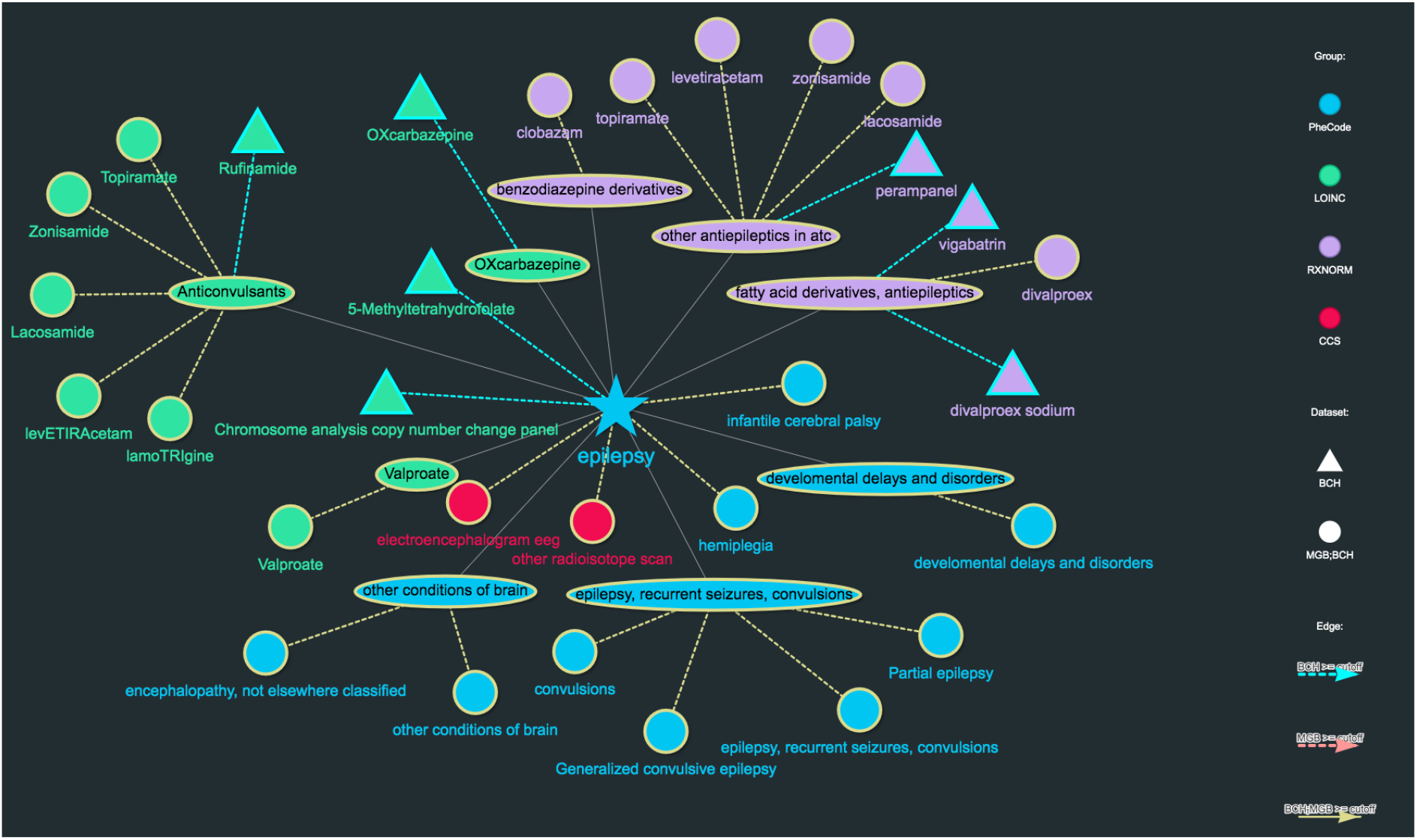
Knowledge graph with top 30 codes selected based on the cosine similarity with Epilepsy using MUGS embeddings for MGB and BCH.

**Figure S2.**
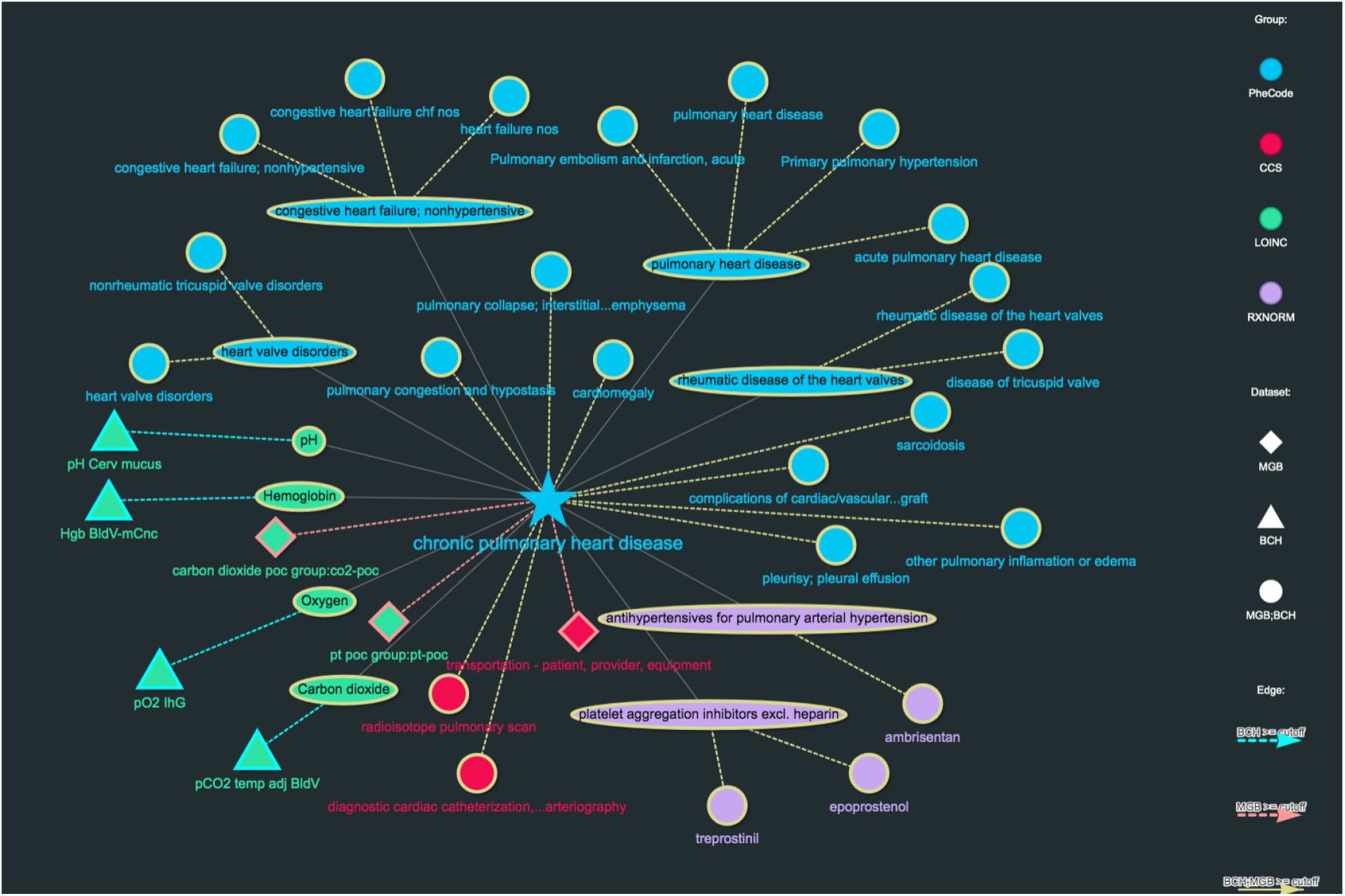
Knowledge graph with top 30 codes selected based on the cosine similarity with PH, also known as chronic pulmonary heart disease, using MUGS embeddings for MGB and BCH.

